# Assessment of fatal cardiovascular disease risk using data-driven diabetes subgroups and SCORE2-Diabetes in 24,943 adults in Mexico City

**DOI:** 10.64898/2025.12.15.25342299

**Authors:** Jerónimo Perezalonso-Espinosa, Juan Pablo Díaz-Sánchez, Daniel Ramírez- García, Karime Berenice Carrillo-Herrera, Leslie Alitzel Cabrera-Quintana, Carlos A. Fermín-Martínez, Martín Roberto Basile-Álvarez, Andrea Malagón-Liceaga, Jaime Berumen, Pablo Kuri-Morales, Roberto Tapia-Conyer, Jesus Alegre-Díaz, Neftali Eduardo Antonio-Villa, Goodarz Danaei, Jacqueline A. Seiglie, Omar Yaxmehen Bello-Chavolla

## Abstract

**BACKGROUND:** Cardiovascular disease (CVD) is a leading cause of diabetes-related mortality in Mexico. Although diabetes subgroups capture underlying disease heterogeneity, their association and utility for risk prediction for fatal CVD in Mexican adults remain unclear.

**METHODS:** We analyzed 24,943 adults with diabetes from the Mexico City Prospective Study. Participants were classified into mild obesity-related (MOD), severe insulin-deficient (SIDD), severe insulin-resistant (SIRD), and mild age-related (MARD) diabetes using a self-normalizing neural network algorithm. Fatal CVD was defined as death from ischemic heart disease or stroke (ICD-10 I20–I25, I60–I69). SCORE2-Diabetes was recalibrated and validated overall and by diabetes subgroup. Cox proportional hazards models were used to estimate subgroup-specific risk, and sequential models evaluated the incremental predictive value of diabetes subgroups combined with SCORE2-Diabetes and traditional risk factors.

**RESULTS:** Over a median follow-up of 19.3 years (IQR 12.7-20.6), 2,223 fatal CVD events (8.9%) were recorded. SIDD was the most prevalent subgroup (50.6%), followed by SIRD (17.3%), MARD (16.8%), and MOD (15.4%). SIDD and MARD showed the highest adjusted risk of fatal CVD (RR 1.58 [95%CI 1.38–1.81] and 1.35 [1.13–1.60]), whereas MOD and SIRD had lower risk. Recalibrated SCORE2-Diabetes demonstrated adequate discrimination overall (c-statistic 0.759, 95%CI 0.745-0.773) and for most subgroups but underperformed in MARD, with recalibration improving risk assessment. The combination of diabetes subgroups and SCORE2-Diabetes improved prediction for fatal CVD outcomes.

**CONCLUSIONS:** Diabetes subgroups show heterogeneity in fatal CVD risk in Mexican adults. SIDD and MARD identify high-risk individuals and integration subgroup classification with SCORE2-Diabetes enhances risk prediction.

## INTRODUCTION

Cardiovascular disease (CVD) is a leading cause of mortality and disability in Mexico^1^. Diabetes mellitus is a key risk factor of CVD, and the importance of multi-risk factor management to reduce CVD burden in this population is well established^2^. However to date few diabetes-specific tools exist to assess CVD risk in this population; moreover, most of these approaches have not been evaluated in populations at high risk of CVD and diabetes-related complications^3,4^. Furthermore, implementation of preventive measures to lower CVD risk amongst individuals living with diabetes in Mexico is currently inadequate^5,6^. Accumulating evidence suggests that early identification of people living with diabetes at high CVD risk could be refined by incorporating factors that consider the heterogeneity of the disease, particularly differences across diabetes phenotypes^7^. Such approaches are especially relevant in contexts like Mexico, where CVD risk factors and mortality cluster in populations with social disadvantage, and where diabetes heterogeneity and its complications remain understudied^8^.

A framework for the classification of data-driven diabetes subgroups derived from clinical parameters has emerged in recent years. This framework identified unique diabetes phenotypes, as well as their association with risk of chronic macro- and microvascular diabetes complications^9,10^. Although these subgroups were validated primarily in European populations, a growing body of research has found their reproducibility in various low- and middle-income countries (LMICs)^11,12^. In Mexico, our group pioneered the development of a framework that reproduced four distinct diabetes subgroups using self-normalizing artificial neural networks (SNNNs): mild obesity-related diabetes (MOD), severe insulin-deficient diabetes (SIDD), severe insulin-resistant diabetes (SIRD), and mild age-related diabetes MARD^13,14^. Currently, there is no evidence on whether each diabetes subgroup carries a differential risk of fatal CVD in Mexican population, or whether CVD risk prediction is improved when diabetes subgroups are considered along with diabetes-specific CVD risk scores. Here, we analyzed data from 24,943 individuals living with diabetes from the Mexico City Prospective Study to assess the risk of fatal CVD risk and the complementary role of CVD risk prediction across diabetes subgroups in Mexican adults.

## METHODS

### Study Population

We analyzed participants enrolled in the Mexico City Prospective Study (MCPS). Full methodological details on recruitment and follow-up have been previously described^15^. MCPS is a blood-based prospective cohort including 159,517 >35 years, recruited by invitation, in the two urban districts of Iztapalapa and Coyoacán in Mexico City between 1998 and 2004. All procedures were approved by Ethics Committees at the Mexican Ministry of Health, the Mexican National Council for Science and Technology, and the University of Oxford, UK. Sociodemographic, health-related, and lifestyle information was collected using an electronic questionnaire. Additionally, all participants had measurements of height, weight, hip circumference (HC), waist circumference (WC), and sitting blood pressure (BP) using calibrated instruments and standard protocols. A non-fasting venous blood sample was obtained, and hemoglobin A1c (HbA1c) levels were measured using a validated high-performance liquid chromatography method10 on HA- 8180 analyzers with calibrators traceable to International Federation of Clinical Chemistry standards. Samples were stored in a central laboratory, separated into plasma and buffy coat, and then frozen to –80°C. From 2018 to 2024, every available plasma sample from the baseline examination was analyzed using nuclear magnetic resonance (NMR) spectroscopy at Nightingale Health Ltd (Kuopio, Finland) and the Clinical Trial Service Unit’s Wolfson Laboratory, quantifying over 249 plasma biomarkers including lipid measures (total cholesterol, HDL-cholesterol, triglycerides and apolipoprotein B).For this analysis, we included individuals with prior self-report of diagnosed diabetes, prior use of glucose-lowering medication, and participants with undiagnosed diabetes but with HbA1c values ≥6.5% at baseline. Age at diabetes diagnosis was estimated as the difference between the half-point of the decade of diabetes diagnosis (ie. 1965 for diagnosis in the 1960 decade) and the year of birth for participants with diagnosed diabetes, and with chronological age for participants with undiagnosed diabetes.

We excluded participants without a report of death status, without decade of diabetes diagnosis, with prior history of cardiovascular disease before cohort entry (defined as angina, non-fatal heart attack, and non-fatal stroke), with BP levels >250 mmHg or <90 mmHg, with a body-mass index (BMI) >80 kg/m2 or <10 kg/m2, or without reported measures of total or HDL cholesterol, or with total cholesterol >20 mmol/l or <1.75 mmol/l (**Figure 1**).

**Figure 1.**
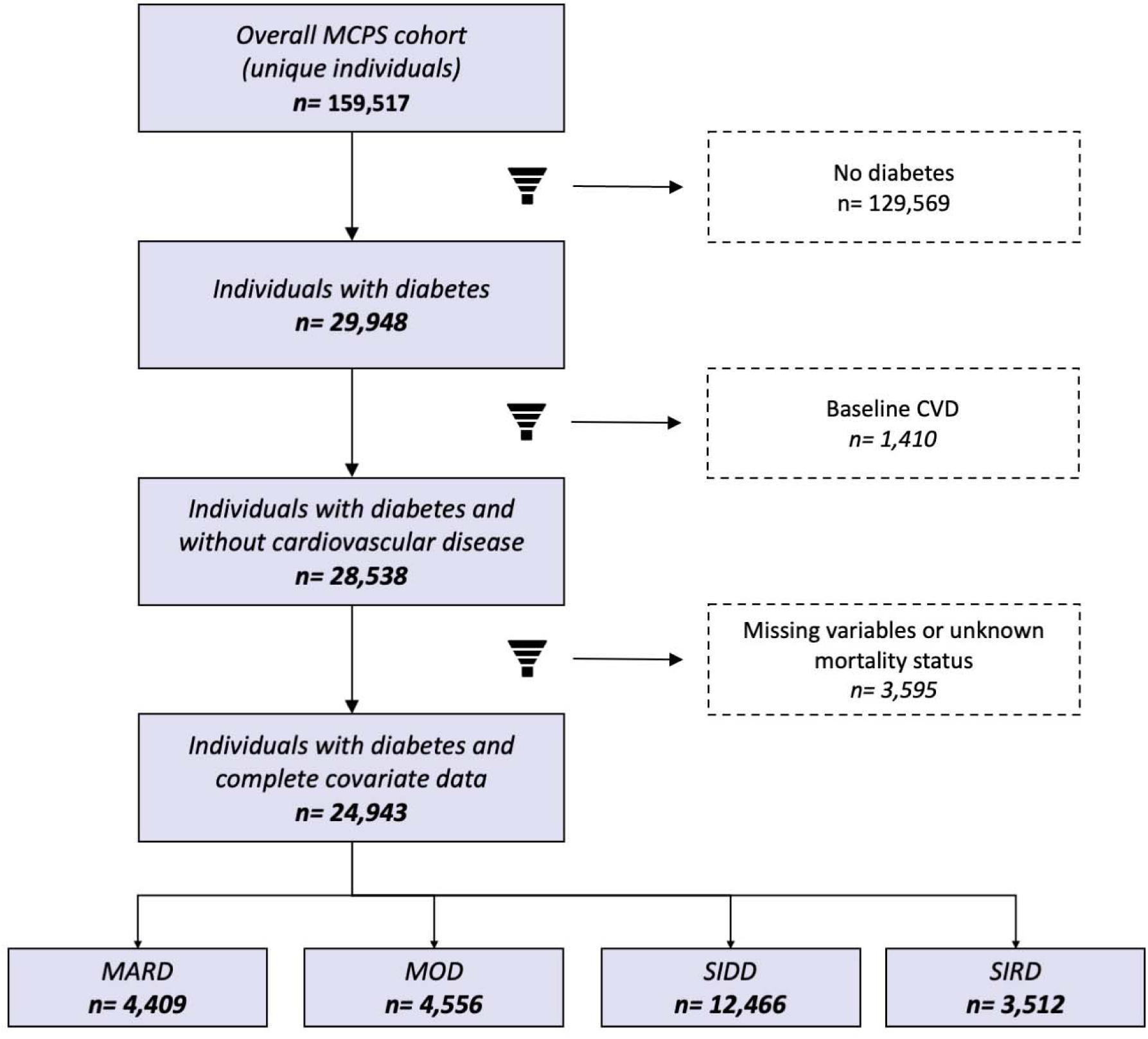
Flowchart of participant selection and exclusion. Flowchart of participant selection and exclusion for the analysis of data-driven diabetes subgroups in the Mexico City Prospective Study.

### Study outcomes

Fatal CVD was defined as death from ischemic heart disease or sudden cardiac death (ICD10 codes I20–I25) or death from stroke (ICD10 codes I60–I69). To identify fatal CVD events, mortality follow-up was done through electronic probabilistic linkage to death registries, with deaths tracked up to September 30^th^, 2022, and contributing causes of death were coded according to the 10th International Classification of Diseases (ICD10). Time to event was evaluated from the date of inclusion into the cohort until the date where the event was reported or censorship at the end of the follow-up time, whichever occurs first.

### Classification of data-driven diabetes subgroups

Diabetes subgroups were classified using SNNNs as previously developed, and applied to U.S. and Mexican populations^16–18^. To apply this model using variables available in MCPS, we adapted our SNNN algorithm by training it in NHANES-III data, and validated it in NHANES-IV compared to k-means clustering results using the original variables proposed by Ahlqvist et al. For the current analyses, the classification considered the following clustering variables: estimated age at diabetes diagnosis, sex, BMI, HbA1c, waist-to-height ratio, systolic and diastolic blood pressure and was implemented using the *keras* R package. This method grouped individuals classified with diabetes into four data-driven phenotypes: MOD, SIDD, SIRD, and MARD. Our algorithm does not explicitly consider severe autoimmune diabetes (SAID) as it requires positive anti-glutamic acid decarboxylase 65 (anti-GAD65) antibodies. Due to the unavailability of these measurements in MCPS, this subgroup was not included in this study. Further details on the development, validation, and performance of this classification algorithm can be consulted in **Supplementary Materials**.

### Statistical analysis

#### Validation of SCORE2-Diabetes for prediction of fatal CVD across diabetes subgroups

CVD risk was assessed using SCORE2-Diabetes^3^, using estimates for a moderate-risk region (100–150 CVD deaths per 100,000 people), based on a reported rate of 146.3 CVD deaths per 100,000 people in Mexico in 2023^19,20^. SCORE2-Diabetes 10-year CVD risk categories were classified as low (<5%), moderate (5 to <10%), high (10 to <20%) and very high risk (≥20%, clinically established atherosclerotic CVD or severe target-organ damage). To validate the use of SCORE2-Diabetes in Mexican population, discrimination for risk of fatal CVD was assessed using Harrell’s c-statistic and time-dependent AUROCs. Calibration of SCORE2-Diabetes was evaluated using mean estimates, slopes, and calibration curves, and overall performance with Brier scores and indices of prediction accuracy (IPAs). These assessments were done for the overall population with diabetes and across data-driven diabetes subgroups. Finally, we recalibrated SCORE2-Diabetes for 10-year fatal CVD outcomes to provide a useful tool for screening adapted to the Mexican context by refitting the linear predictor of SCORE2-Diabetes to re-estimate the calibration slope at 10 years and baseline survival where the linear predictor equals to zero. Recalibrated individual risks were obtained by applying the fitted slope and baseline risk to each person’s linear predictor and then transforming back to fatal CVD risk, as detailed in **Supplementary Materials**

#### Assessment of fatal CVD risk across diabetes subgroups

Kaplan-Meier estimators with inverse probability weighting (IPW) were used to estimate cumulative incidence of fatal CVD across diabetes subgroups, with IPW adjustment for age, sex, duration of diabetes, diabetes treatment, antihypertensive treatment, statin treatment, number of comorbidities, educational level, smoking, and socioeconomic status.

Adjusted rates of overall, cerebrovascular, and ischemic cardiac fatal events were presented as cumulative incident rates and were plotted for each outcome separately. Next, we estimated cause-specific mortality rate ratios (RR) using Cox proportional hazard regression models adjusted for sex, years since diagnosis, diabetes treatment, number of comorbidities, educational attainment, municipality, smoking status, social development index and physical activity and stratified for age-at-risk per five-year increments using a Lexis expansion.

#### Diabetes subgroups and SCORE2-Diabetes for prediction of fatal CVD

Next, we evaluated the complementary role of diabetes subgroups and SCORE2-Diabetes for prediction of fatal CVD risk using sequential Cox proportional hazards models as follows: 1) Model 1 included sex, age, years since diagnosis, diabetes treatment, number of comorbidities, educational attainment, municipality, smoking status, social development index and physical activity. 2) Model 2 included variables in Model 1 along with included diabetes subgroups, using MOD as the reference subgroup 3) Model 3 included variables in Model 1 along with SCORE2-Diabetes, and 4) Model 4 combined Model 1 with both diabetes subgroups and SCORE2-Diabetes. Models were compared using Harrel’s c-statistics, and changes in the Akaike Information Criteria compared to Model 1 for prediction of fatal CVD. All statistical analyses were conducted using R software version 4.5.2.

## RESULTS

### Study population

Among 159,517 participants enrolled in MCPS, 29,948 participants had diabetes at baseline (18.77%); amongst these, 1,410 participants who had self-reported prior diagnosis of CVD at baseline (4.7%) were excluded.. Also, 3,595 participants with missing mortality status or covariate data for cluster classification or SCORE2-Diabetes calculation were excluded from the study; therefore, we included only24,943 adults with diabetes at baseline (**Figure 1**). Participants with diabetes had a mean age of 58±12 years with a mean follow-up of 16±6 years, were predominantly female (66.7%), and amongst other measures they had mean BMI of 29.7±5.4 kg/m^2^, systolic BP of 134±18 mmHg, and HbA1c of 8.86±2.37% (**Table 1**). SIDD was the most prevalent diabetes subgroup (50.6%), had higher HbA1c levels and higher rate of insulin use compared to other subgroups. SIRD accounted for 17.3% of participants and was characterized by a high BMI and higher METS-IR, followed by MARD (16.8%) whom had an older age at diagnosis. Finally, 15.4% of participants were in the MOD subgroup, which had mild alterations in anthropometric and metabolic measures (**Table 1**).

**Table 1.** Characteristics of adult-onset diabetes subgroups in the Mexico City Prospective Study. Characteristics of the general diabetes population in the Mexico City Prospective Study and across diabetes subgroups. **Abbreviations:** SIRD: severe insulin-resistant diabetes; SIDD: severe insulin-deficient diabetes; MARD: mild age-related diabetes; MOD: mild obesity-related diabetes. Clusters based on Bello-Chavolla et. al. (2020) & Ahlqvist et. al. (2018). METS-IR: Metabolic Score for Insulin Resistance; eGFR: Estimated glomerular filtration rate.

| Variable | All<br>n = 24,943<br>(100%) | SIDD<br>n = 12,466<br>(50%) | MOD<br>n = 4,556<br>(18.3%) | MARD<br>n = 4,409<br>(17.7%) | SIRD<br>n = 3,512<br>(14.1%) |
| --- | --- | --- | --- | --- | --- |
| Age at recruitment (years) | 58 (12) | 56 (11) | 53 (10) | 71 (8) | 55 (10) |
| Male (%) | 8,188 (33%) | 4,209 (34%) | 1,712 (38%) | 1,624 (37%) | 643 (18%) |
| Estimated age at diabetes<br>diagnosis (years) | 52 (12) | 49 (11) | 45 (8) | 68 (8) | 52 (9) |
| Undiagnosed diabetes (%) | 7,041 (28%) | 2,662 (21%) | 1,023 (22%) | 1,690 (38%) | 1,666 (47%) |
| Smoking (%) | 6,539 (26%) | 3,521 (28%) | 1,379 (30%) | 790 (18%) | 849 (24%) |
| Body mass index (kg/m<sup>2</sup>) | 29.7 (5.4) | 28.5 (4.6) | 27.9 (3.4) | 28.1 (3.5) | 38.0 (4.9) |
| Systolic blood pressure<br>(mmHg) | 134 (18) | 133 (18) | 130 (18) | 139 (18) | 137 (17) |
| Diastolic blood pressure<br>(mmHg) | 85 (10) | 85 (10) | 83 (10) | 86 (11) | 88 (10) |
| Waist-To-Hip Ratio | 0.93 (0.07) | 0.93 (0.07) | 0.92 (0.07) | 0.94 (0.07) | 0.92 (0.08) |
| Waist-To-Height Ratio | 0.64 (0.08) | 0.62 (0.08) | 0.60 (0.06) | 0.64 (0.07) | 0.73 (0.07) |
| Glucose (mg/dl) | 207 (117) | 282 (111) | 121 (56) | 134 (58) | 143 (64) |
| Glycated HbA1c (%) | 8.86 (2.37) | 10.79 (1.70) | 6.56 (0.82) | 7.01 (0.86) | 7.28 (1.01) |
| METS-IR (unit or %) | 51 (10) | 51 (9) | 46 (7) | 47 (8) | 64 (9) |
| Missing | 6 (<0.1%) |  | 3 (<0.1%) | 1 (<0.1%) | 2 (<0.1%) |
| Creatinine (mg/dl) | 0.80 (0.56) | 0.75 (0.34) | 0.96 (1.06) | 0.82 (0.37) | 0.74 (0.40) |
| eGFR (ml/min/1.73m<sup>2</sup>) | 96 (20) | 99 (18) | 95 (25) | 86 (17) | 98 (18) |
| Total cholesterol (mg/dl) | 169 (40) | 173 (41) | 165 (40) | 163 (38) | 166 (37) |

| **Variable** | **All**<br>**n = 24,943**<br>**(100%)** | **SIDD**<br>**n = 12,466**<br>**(50%)** | **MOD**<br>**n = 4,556**<br>**(18.3%)** | **MARD**<br>**n = 4,409**<br>**(17.7%)** | **SIRD**<br>**n = 3,512**<br>**(14.1%)** |
| --- | --- | --- | --- | --- | --- |
| HDL cholesterol (mg/dl) | 39 (8) | 40 (8) | 38 (8) | 38 (8) | 38 (7) |
| Non-HDL cholesterol (mg/dl) | 130 (37) | 133 (38) | 127 (36) | 125 (34) | 128 (34) |
| Total triglycerides (mg/dl) | 164 (70) | 175 (75) | 151 (65) | 150 (60) | 158 (64) |
| Apolipoprotein B (mg/dl) | 92 (23) | 94 (24) | 90 (23) | 89 (21) | 91 (22) |
| Metformin (%) | 3,252 (13%) | 1,942 (16%) | 567 (12%) | 412 (9.3%) | 331 (9.4%) |
| Sulphonylurea (%) | 12,428 (50%) | 7,078 (57%) | 2,171 (48%) | 1,906 (43%) | 1,273 (36%) |
| Insulin (%) | 1,320 (5.3%) | 959 (7.7%) | 195 (4.3%) | 78 (1.8%) | 88 (2.5%) |
| Other antidiabetic treatments (%) | 258 (1.0%) | 145 (1.2%) | 50 (1.1%) | 40 (0.9%) | 23 (0.7%) |

### Cardiovascular events and CVD risk across diabetes subgroups

A total of 2,218 fatal CVD events (8.9%) were recorded among participants with diabetes up to September 30^th^, 2022. Most events were fatal ischemic cardiac events (7.4%), and a minority were fatal cerebrovascular events (1.5%). Among subgroups, MARD had the highest proportion of fatal CVD events (12%), followed by SIDD (9.6%). SIRD and MOD had a similar percentage of events with 6.2% and 6.1%, respectively (**Table 2**). Using IPW-adjusted Kaplan-Meier estimators, we observed the highest adjusted fatal CVD rate for SIDD and MARD; on the other hand, participants in the MOD subgroup had the lowest adjusted fatal CVD rate, followed by SIRD (**Figure 2**). When fitting adjusted Cox proportional hazards model with the Lexis expansion and using MOD as the reference subgroup, SIDD showed a significantly higher risk of fatal CVD (RR 1.58, 95%CI 1.38– 1.81), as did MARD (RR 1.35, 95%CI 1.13-1.60), with no significant differences with the SIRD subgroup (**Table 3** and **Figure 2**).

**Figure 2.**
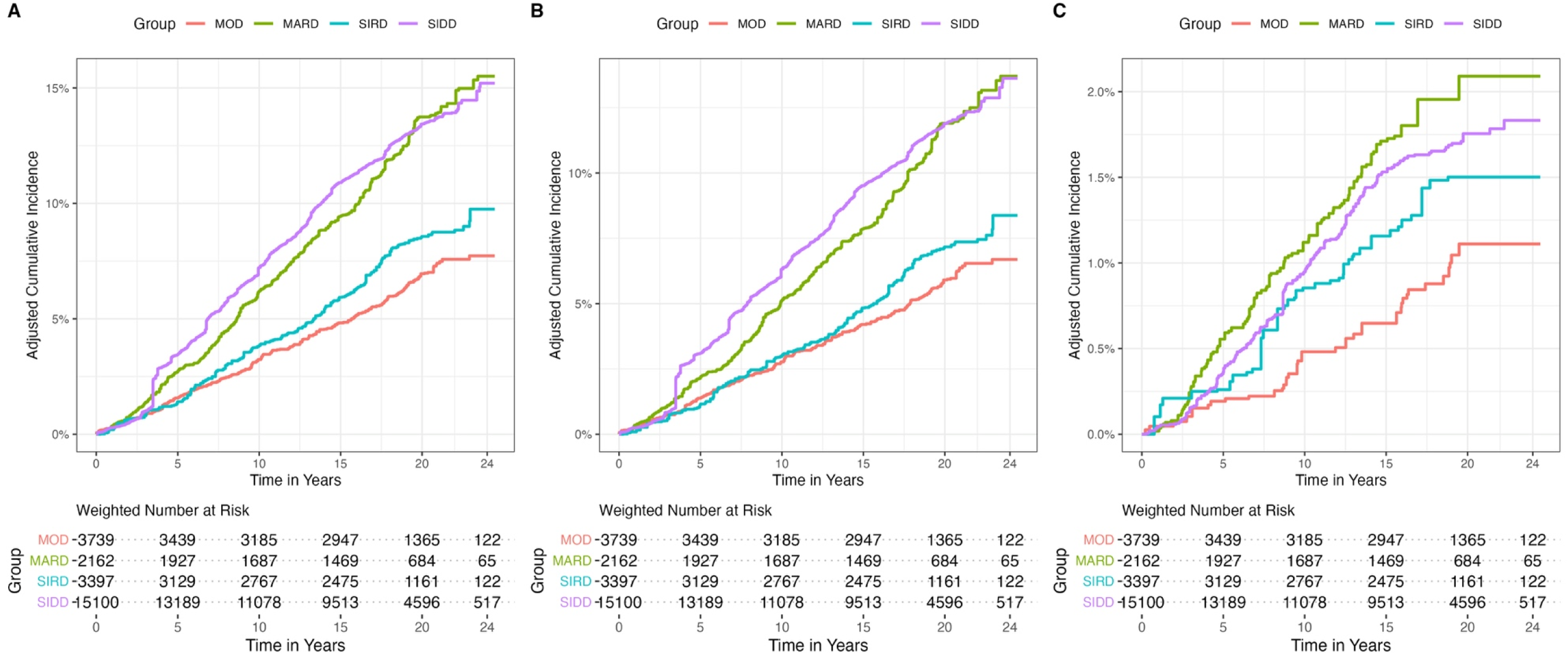
Kaplan-Meier curves for fatal cardiovascular disease events across diabetes subgroups in the Mexico City Prospective Study. Kaplan-Meier estimates with Inverse Probability Weighting (IPW) adjustment for age, sex, duration of diabetes, diabetes treatment, antihypertensive treatment, statin treatment, number of comorbidities, educational level, smoking, total cholesterol, triglyceride levels and socioeconomic status for diabetes subgroups over the total follow-up period showing risk of overall fatal CVD (**A**), fatal myocardial infarction (**B**) and fatal stroke (**C**).

**Table 2.** Fatal cardiovascular events among diabetes subgroups in the Mexico City Prospective Study Cohort. Total number and cumulative incident rates of fatal cardiovascular events among the general diabetes population in MCPS and diabetes subgroups up to September 30^th^, 2022. **Abbreviations:** SIRD: severe insulin-resistant diabetes; SIDD: severe insulin-deficient diabetes; MARD: mild age-related diabetes; MOD: mild obesity-related diabetes. Clusters based on Bello-Chavolla et. al. (2020) & Ahlqvist et. al. (2018).

|  | Overall diabetes | SIDD | MOD | MARD | SIRD |
| --- | --- | --- | --- | --- | --- |
| Variable | n = 24,943 | n = 12,466 | n = 4,556 | n = 4,409 | n = 3,512 |
| Total cardiovascular deaths<sup>1</sup> | 2,218 (8.9%) | 1,178 (9.4%) | 276 (6.1%) | 563 (13%) | 201 (5.7%) |
| Ischemic cardiac deaths | 1,850 (7.4%) | 990 (7.9%) | 238 (5.2%) | 459 (10%) | 163 (4.6%) |
| Cerebrovascular deaths | 368 (1.5%) | 188 (1.5%) | 38 (0.8%) | 104 (2.4%) | 38 (1.1%) |

**Table 3.** Hazard Ratios for the Lexis-expanded Cox model. Hazard ratios for Lexis-expanded Cox model stratified for age-at-risk per 5-year increments. **Abbreviations:** CI: Confidence Interval, HR: Hazard Ratio, SIRD: severe insulin-resistant diabetes; SIDD: severe insulin-deficient diabetes; MARD: mild age-related diabetes; MOD: mild obesity-related diabetes.

| Diabetes subgroup | Cardiovascular death |  | Ischemic cardiac death |  | Cerebrovascular death |  |
| --- | --- | --- | --- | --- | --- | --- |
|  | HR | 95% CI | HR | 95% CI | HR | 95% CI |
| MOD | — | — | — | — | — | — |
| MARD | 1.35 | 1.14, 1.61 | 1.31 | 1.09, 1.58 | 1.60 | 1.02, 2.50 |
| SIRD | 1.04 | 0.86, 1.26 | 1.01 | 0.82, 1.24 | 1.22 | 0.76, 1.98 |
| SIDD | 1.58 | 1.38, 1.81 | 1.54 | 1.33, 1.79 | 1.82 | 1.26, 2.62 |

**Note:** Models were adjusted for covariates measured at baseline, including sex, estimated years since diabetes diagnosis, diabetes, hypertension and statin treatment, number of comorbidities, educational attainment, municipality of residence, smoking status, social development index, physical activity, total cholesterol, and total triglycerides.

### Validation of SCORE2-Diabetes for prediction of fatal CVD risk

Using the original SCORE2-Diabetes algorithm, we identified that most adults with diabetes were at high 10-year risk of CVD (9,019/24,943, 36.2%), followed by moderate risk (6,951/24,943, 27.9%), very high risk (4,878/24,943, 19.6%), and low risk (4,065/24,943, 16.3%, **Supplementary Figure 1**). Regarding diabetes subgroups, MARD and SIDD were had the highest level of cardiovascular risk, where 83.3% of participants in the MARD subgroup were in the high or very high-risk category and only 1% were considered low risk. Similarly, most of SIDD participants were in the two highest risk categories. In SIRD, 38% were in the moderate category and 32% in the low-risk category. Finally, MOD had the biggest proportion of participants in the low-risk category (37%, **Supplementary Figure 1**). When evaluating the discrimination of SCORE2-Diabetes for prediction of 10-year risk of fatal CVD, the highest performance was observed in the MOD subgroup (Harrel’s C= 0.804, 95%CI 0.769-0.389), followed by SIRD (0.794, 95%CI 0.743-0.845) and SIDD (0.736, 95%CI 0.716-0.756). These subgroups demonstrated discrimination similar to that of the overall population with diabetes, whereas markedly lower discrimination was observed for the MARD subgroup (0.682, 95%CI 0.655-0.709). Calibration analysis revealed that original SCORE2-Diabetes algorithm results in a systematic and substantial overprediction, and it was more accurate in the more severe diabetes phenotypes and poorest in MARD (**Table 4**). Recalibration of SCORE2-Diabetes improved agreement between predicted and observed 10-year risk of fatal CVD outcomes for Mexican adults with diabetes with slight decreased on discrimination measures overall and for most diabetes subtypes; however, discrimination and calibration remained lower for MARD using the recalibrated SCORE2-Diabetes (**Table 4**). When assessing risk categories using the recalibrated SCORE2-Diabetes for fatal CVD outcomes, most adults with diabetes were now at lower risk categories, whilst MARD and SIDD remained as the subgroups with most participants at high- and very high-risk of fatal CVD outcomes (**Supplementary Figure 2**).

**Table 4.** Performance of SCORE2-Diabetes for 10-year risk of fatal cardiovascular disease amongst 24,943 adults with diabetes and without cardiovascular disease enrolled in the Mexico City Prospective Study overall and stratified by diabetes subgroup. Abbreviations: IPA: Index of Prediction Accuracy, SIRD: severe insulin-resistant diabetes; SIDD: severe insulin-deficient diabetes; MARD: mild age-related diabetes; MOD: mild obesity-related diabetes. Clusters based on Bello-Chavolla et. al. (2020) & Ahlqvist et. al. (2018).

| SCORE2-Diabetes | Diabetes Subgroup | c-statistic (95%CI) | Predicted risk | Observed risk | O/E ratio (95%CI) | Calibration slope (95%CI) | Brier score | IPA (%) |
| --- | --- | --- | --- | --- | --- | --- | --- | --- |
| Original | Overall | 0.759 | 0.222 | 0.048 | 0.216 | 0.630 | 0.040 | 8.72 |
|  |  | (0.745-0.773) |  |  | (0.203 - 1.086) | (0.587 - 0.673) | (0.037-0.042) |  |
|  | MOD | 0.804<br>(0.769-0.839) | 0.147 | 0.034 | 0.227<br>(0.193 - 0.268) | 0.663<br>(0.563 - 0.762) | 0.039<br>(0.037-0.042) | 9.65 |
|  | MARD | 0.682<br>(0.655-0.709) | 0.407 | 0.078 | 0.192<br>(0.172 - 0.215) | 0.584<br>(0.482 - 0.685) | 0.040<br>(0.038-0.043) | 7.35 |
|  | SIRD | 0.794<br>(0.743-0.845) | 0.124 | 0.025 | 0.200<br>(0.161 - 0.248) | 0.784<br>(0.624 - 0.945) | 0.039<br>(0.037-0.041) | 9.83 |
|  | SIDD | 0.736<br>(0.716-0.756) | 0.211 | 0.050 | 0.235<br>(0.216 - 0.255) | 0.621<br>(0.557 - 0.685) | 0.040<br>(0.037-0.042) | 9.00 |
| Recalibrated | Overall | 0.748<br>(0.734-0.762) | 0.051 | 0.048 | 0.933<br>(0.88 - 1.086) | 1.0<br>(0.930 - 1.070) | 0.040<br>(0.038-0.042) | 7.63 |
|  | MOD | 0.799<br>(0.764-0.834) | 0.036 | 0.034 | 0.919<br>(0.846 - 0.997) | 1.096<br>(0.928 - 1.263) | 0.040<br>(0.037-0.042) | 9.01 |
|  | MARD | 0.662<br>(0.633-0.691) | 0.086 | 0.078 | 0.908<br>(0.732 - 1.126) | 0.833<br>(0.673 - 0.993) | 0.040<br>(0.038-0.043) | 6.83 |
|  | SIRD | 0.79<br>(0.741-0.839) | 0.034 | 0.025 | 0.722<br>(0.646 - 0.808) | 1.287<br>(1.019 - 1.554) | 0.040<br>(0.037-0.042) | 8.68 |
|  | SIDD | 0.724<br>(0.704-0.744) | 0.049 | 0.050 | 1.009<br>(0.856 - 1.19) | 0.990<br>(0.883 - 1.096) | 0.040<br>(0.038-0.043) | 6.75 |

### Predictors of CVD risk on adults with diabetes

Amongst all evaluated models, the best-performing was Model 4, which combined diabetes-specific risk factors, diabetes subgroups and the recalibrated SCORE2-Diabetes (c-statistic 0.725, 95%CI 0.715-0.735), followed closely by Model 2 (Risk factors + diabetes subgroups; c-statistic 0.725, 95%CI 0.715-0.735), and Model 3 (Risk factors + recalibrated SCORE2-Diabetes; c-statistic 0.720, 95%CI 0.710-0.731). Compared to Model 1 (Risk factors), Model 2 had larger decreases in AIC compared to Model 3, indicating improved predictive performance by adding diabetes subgroups. Moreover, Model 4 (Risk factors + SCORE2-Diabetes + Diabetes subgroups) showed the largest decrease in AIC compared to Model 1. These trends were similar when analyzing ischemic cardiac and cerebrovascular deaths separately (**Figure 3**).

**Figure 3.**
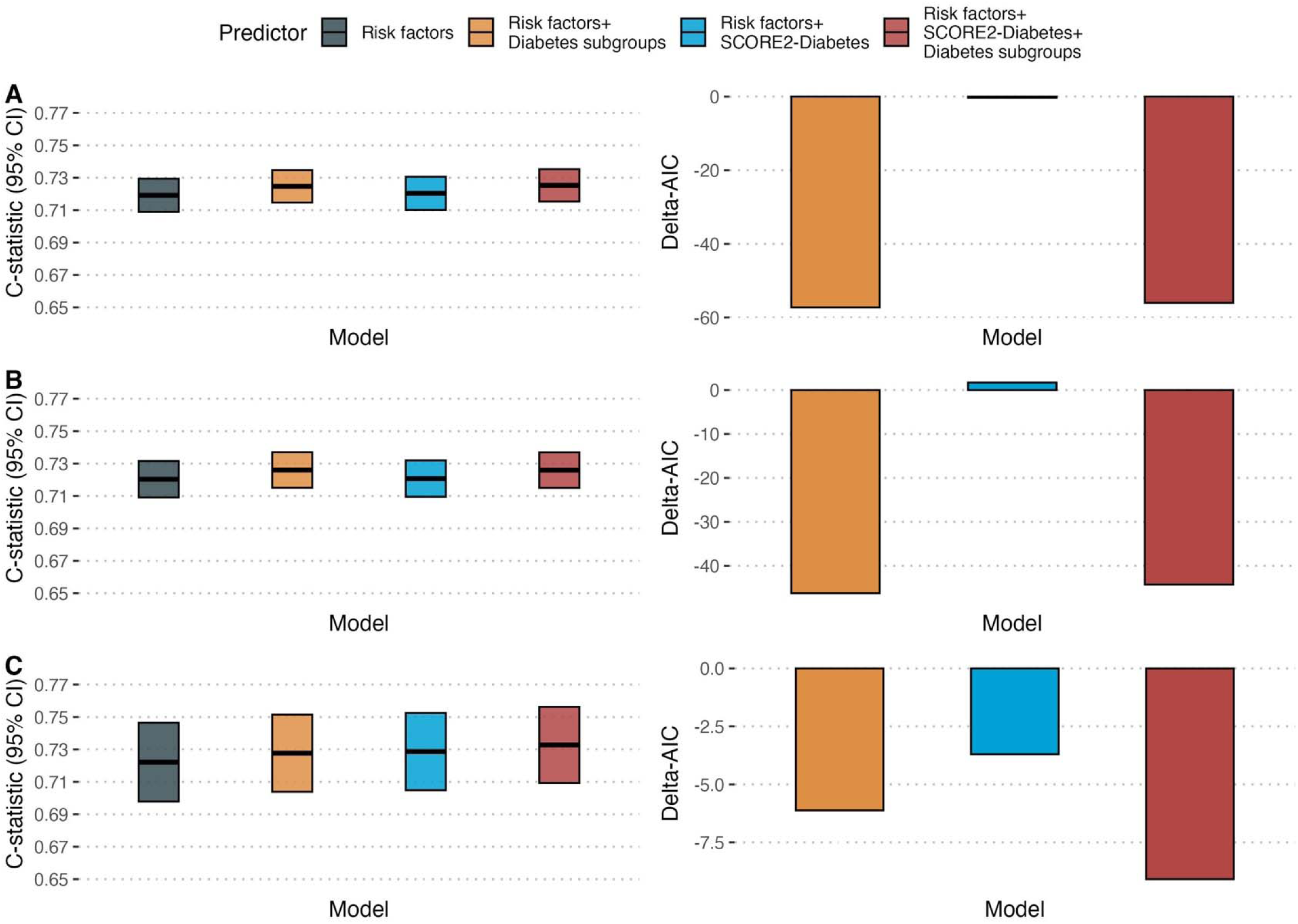
Model performance for prediction of fatal cardiovascular outcomes in the Mexico City Prospective Study. Harrel’s c-statistic and 95% confidence intervals, as well as Differences in the Akaike Information Criteria (Delta-AIC) for the four Cox proportional hazards models for overall cardiovascular (A), ischemic cardiac (B), and cerebrovascular deaths (C). Model 1 included sex, age, years since diagnosis, diabetes treatment, number of comorbidities, educational attainment, municipality, smoking status, social development index, physical activity, total cholesterol and total triglycerides. Model 2 included variables in Model 1 along with included diabetes subgroups, using MOD as the reference subgroup 3) Model 3 combined Model 1 with the recalibrated SCORE2-Diabetes, and 4) Model 4 combined Model 1 with both diabetes subgroups and the recalibrated SCORE2-Diabetes. The ΔAIC was calculated by subtracting the AIC of model 1 from that of the other three models.

## DISCUSSION

In this large cohort of adults living with diabetes in Mexico City, we observed substantial heterogeneity in long-term risk of fatal CVD. Consistent with previous reports, the SIDD phenotype was the most prevalent in MCPS, followed by MOD, MARD, and SIRD, a distribution that differs from patterns reported in European and U.S. populations, where MARD is the predominant diabetes subtype^17,18^. These subgroups exhibited marked differences in cardiovascular outcomes, where SIDD and MARD had the highest rates of fatal CVD, including ischemic heart and cerebrovascular deaths, whereas MOD and SIRD were associated with substantially lower risk. Risk stratification using the recalibrated SCORE2-Diabetes closely aligned with observed outcomes, and IPW-adjusted analyses identified SIDD and MARD as the subgroups at greatest risk of fatal CVD, highlighting the contributions of insulin deficiency and aging as key determinants of CVD mortality in Mexican adults with diabetes. SCORE2-Diabetes demonstrated adequate discrimination across most subgroups, with the exception of MARD, suggesting that existing risk algorithms may be less precise for this phenotype, which is the most prevalent in U.S. and European populations^18,21^. Notably, most cardiovascular risk algorithms limit inclusion of participants ≥80 years for their development; specifically, SCORE2 (which is the basis for risk estimation in SCORE2-Diabetes) primarily included participants aged 40-79 years and is recommended to be used in participants 40-69 years which exclude most participants who would be classified as MARD. Predictive modeling further indicated that incorporation of diabetes subgroup classification improved prediction of fatal CVD beyond traditional risk factors alone and enhanced the performance of the recalibratedSCORE2-Diabetes when combined with diabetes-specific variables. Collectively, these findings underscore the clinical and public health importance of accounting for diabetes heterogeneity when assessing CVD risk. Integrating subgroup classification with established risk scores and diabetes-specific factors may facilitate more precise risk stratification and support the development of tailored preventive strategies to reduce CVD mortality in this population.

A key finding of our study is the marked variation in fatal CVD events across diabetes subgroups. Prior studies examining CVD outcomes by diabetes subgroup have reported inconsistent results, likely reflecting shorter follow-up periods and smaller sample sizes^22–24^. Our results indicate that the SIDD and MARD phenotypes had the highest adjusted risk of fatal CVD, with lower rates observed for the MOD and SIRD phenotypes. This is relevant given that SIDD and MARD are the most prevalent diabetes subgroups across many regions. Studies from Germany, Japan, the UK, China, Norway, and the US similarly have identified MARD as the most prevalent diabetes subgroup^22,23,25–28^. In contrast, a prior nationally representative study of Mexican adults conducted by our group identified SIDD as the most prevalent subgroup, particularly in Southern Mexico^17^. In line with these findings, in MCPS, severe diabetes phenotypes (SIDD and SIRD) accounted for over two-thirds of participants with diabetes, while milder phenotypes (MOD and MARD) were less common. Given that SIDD displayed the highest risk of fatal CVD in MCPS it is likely that insulin deficiency, likely related to poor glycemic control, is a main driver of CVD among Mexican adults with diabetes. Future research should examine whether specific preventive strategies, including lipid-lowering intensification, tighter glycemic control, and the use of cardioprotective medications (eg. SGLT-2 inhibitors and GLP-1 receptor agonists) could reduce CVD risk for specific diabetes subgroups^29^.

Our study also provides the first external validation of SCORE2-Diabetes for prediction of 10-year fatal CVD risk in Mexican population, and the first evaluation of its performance across distinct diabetes subgroups in any population. Although multiple CVD risk calculators exist, SCORE2-Diabetes is among the few specifically developed for individuals with diabetes and validated in large, diverse cohorts^30,31^. Overall, SCORE2- Diabetes demonstrated adequate discrimination for fatal CVD overall and for most diabetes subgroups, with notably lower performance for the MARD phenotype. This is of concern, as MARD exhibited higher fatal CVD risk than both MOD and SIRD in our cohort, indicating a need for improved risk estimation for this subgroup. Calibration analyses revealed systematic overprediction, which is expected given that the original SCORE2- Diabetes was originally developed for both fatal and non-fatal outcomes^3^. Despite this, risk estimates and category assignments using the recalibrated SCORE2-Diabetes in MCPS aligned closely with observed fatal CVD events across all diabetes subgroups. The recalibration of SCORE2-Diabetes for fatal outcomes may be a useful tool for assessment of fatal CVD risk in a Mexican context. However, additional external validation and recalibration of SCORE2-Diabetes for both fatal and non-fatal outcomes overall and across diabetes subgroups is still needed to fully establish the clinical utility of this score for Mexican adults with diabetes.

After establishing differences in fatal CVD risk across diabetes subgroups, we evaluated the predictive performance of subgroups alone and in combination with other relevant risk factors and SCORE2-Diabetes. We observed that the addition of diabetes subgroups to diabetes-specific risk factors increased the predictive performance for fatal CVD outcomes and provided larger improvements compared to the sole addition of the recalibrated version of SCORE-2-Diabetes. Strikingly, the simultaneous addition of the recalibrated version of SCORE2-Diabetes and diabetes subgroups yielded the better model, likely indicating that consideration of diabetes heterogeneity likely enhances CVD risk prediction. Our approach represents an initial step toward incorporating data-driven diabetes subgroups into more complex predictive frameworks and shows its complementary role to diabetes-specific CVD risk models. Further research is needed to explore whether incorporating diabetes subgroups could yield more robust and accurate CVD risk models to improve estimation of CVD risk in diabetes.

Our study has several strengths. Unlike prior studies of fatal CVD outcomes in diabetes subgroups which were often limited by small sample sizes, our study leverages one of the largest cohorts of adults with diabetes in Mexico and benefits from long-term follow-up aligned with standard CVD risk prediction horizons. To our knowledge, ours is the first study to characterize differences in fatal CVD risk across diabetes subgroups and to externally validate SCORE2-Diabetes in Mexican adults. However, some limitations should be considered when interpreting our findings. First, MCPS is not nationally representative and is restricted to an urban population; therefore, our results may not generalize to adults with diabetes living in rural areas. Second, the lack of consistent assessment of non-fatal CVD events in MCPS means that our analysis is limited to fatal outcomes and may not capture the full spectrum of CVD risk in individuals with diabetes and limits our ability to adequately assess the calibration of SCORE2-Diabetes in our cohort; furthermore, SCORE2-Diabetes also includes outcomes related to hear failure, arrythmias and aortic aneurisms, which were not included in our definition and thus may influence their performance. Third, previous research has shown that the phenotype of diabetes subgroups is not necessarily stable over time, and migration between subgroups was not accounted for when assessing the risk of fatal CVD outcomes. Finally, our analysis does not reflect changes in diabetes management over the past two decades, including the introduction of cardioprotective therapies such as GLP-1 receptor agonists and SGLT2 inhibitors, nor does it differentiate participants who achieved glycemic targets over time from those who did not.

## Conclusions

In this large prospective cohort of Mexican adults with diabetes, we identified substantial heterogeneity in clinical characteristics and long-term fatal CVD risk which was effectively captured using data-driven diabetes subgroups. The SIDD and MARD phenotypes were associated with the highest rates of fatal CVD, underscoring the central role of insulin deficiency and older age as determinants of cardiovascular mortality in adults with diabetes in Mexico. Integrating diabetes subgroups with diabetes-specific risk factors improved predictive performance for fatal CVD risk, outperforming and complementing the incorporation of SCORE2-Diabetes into this framework. We also performed an external validation and recalibration of SCORE2-Diabetes for fatal CVD outcomes, which may provide a useful tool for risk stratification in the Mexican context. Our findings highlight the clinical and public health value of integrating diabetes subgroup classification into CVD risk assessment and prevention strategies, particularly for high-risk phenotypes, and provide a foundation for refining predictive models and developing targeted interventions to reduce CVD mortality in diverse populations with diabetes.

## ETHICS APPROVAL AND CONSENT TO PARTICIPATE

The study was approved by Ethics Committees at the Mexican Ministry of Health, the Mexican National Council for Science and Technology, and the University of Oxford, UK. All participants provided written informed consent.

## CONSENT FOR PUBLICATION

Not applicable

## AVAILABILITY OF DATA AND MATERIALS

Data from the Mexico City Prospective Study are available to bona fide researchers. For more details, the study’s Data and Sample Sharing policy may be downloaded (in English or Spanish) from https://www.ctsu.ox.ac.uk/research/mcps. Available study data can be examined in detail through the study’s Data Showcase, available at https://datashare.ndph.ox.ac.uk/mexico/. Code and Supplementary Materials and Methods are available for reproducibility of results at https://github.com/oyaxbell/cvd_risk_diabetes_mcps.

## AUTHOR CONTRIBUTIONS

Establishing the cohort: JBC, PKM, JAD and RTC. Obtaining funding: JBC, PKM, JAD, RTC and OYBC. Data acquisition, analysis, or interpretation of data: JPE, DRG, JPDS, KBCH, LACQ, CAFM, AML, MRBA, JBC, PKM, RTC, JAD, JAS, DG, NEAV. Drafting first version of manuscript: JPE, OYBC. Critical revision of the report for important intellectual content: All authors. All authors have seen and approved the final version and agreed to its publication. Each author contributed important intellectual content during manuscript drafting or revision and accepts accountability for the overall work by ensuring that questions pertaining to the accuracy or integrity of any portion of the work are appropriately investigated and resolved.

## Data Availability

https://github.com/oyaxbell/cvd_risk_diabetes_mcps

## ACKNOWLEDGMENTS

This project was registered and approved by the Research Committee at Instituto Nacional de Geriatría, project number DI-PI-009/2024. JPE, CAFM, and JPDS are enrolled at the PECEM Program of the Faculty of Medicine at UNAM and are supported by SECIHTI. JAS was supported by Grant Number K23DK135798 from the NIH/NIDDK and by the Massachusetts General Hospital Executive Committee and Center for Diversity and Inclusion Physician-Scientist Development Award. The authors thank the participants for their willingness to take part in this prospective study 20 years ago. This research was conducted using Mexico City Prospective Study (MCPS) data obtained through an open-access data request (application number 2022-012).

