## Supplementary Material for "Assessment of fatal cardiovascular disease risk using data-driven diabetes subgroups and SCORE2-Diabetes in 24,943 adults in Mexico City"

### SUPPLEMENTARY METHODS

*Adaptation of the SNNN algorithm to MCPS data*

***Cluster classification in NHANES-III and NHANES-IV***

Based on methods previously developed by our group[1], we adapted our self-normalizing artificial neural network (SNNN) algorithm to fit MCPS data using NHANESS-III as a training dataset and NHANES-IV as a validation dataset. Amongst 13,866 participants in NHANES-III, 1,665 had diabetes defined as self-reported diabetes, HbA1c ≥6.5%, or fasting plasma glucose ≥126mg/dL; similarly, amongst 41,810 participants in NHANES-IV 6,392 had diabetes as per the same definition. Next, we included only participants with recently-diagnosed diabetes defined as ≤5 years from diabetes onset, and filtered those with HOMA2-IR, HOMA2-B, age from diabetes onset, BMI and HbA1c ≥5 standard deviations from the population mean to derive diabetes clusters as proposed by Ahlqvist et al.[2], yielding 1,017 participants in NHANES-III and 449 participants in NHANES-IV with complete data to estimate diabetes clusters. In both NHANES-III and NHANES-IV, diabetes subgroups were classified using k-means clustering after data centering and scaling to mean 0 and standard deviation of 1, to derive a four cluster solution with 1,000 runs using the *fpc* R package and assessed cluster stability using clusterwise Jaccard jittering means yielding coefficients >0.97 using 1,000 boostrap samples with the *clusterboot* R package.

***Fitting of the modified SNNN algorithm***

We used the derived cluster classification in NHANES-III as the gold standard and developed a SNNN algorithm using the *keras* R package, which classified diabetes subgroups based on variables available in the MCPS dataset including BMI, systolic and diastolic blood pressure, HbA1c, age at diabetes diagnosis and the waist-to-height ratio (WHtR). The SNNN model had three hidden layers, including a 32-unit dense layer with a LeCun normal intializer, an alpha droput layer with an alpha rate of 0.2 and a 4-unit dense layer with a Softmax activation function as the output layer. The model was fitted using an ADAM optimizer, a SeLU activation function, and a sparse categorical cross-entropy loss function, and was trained for 20 epochs using the *keras* and *reticulate* R package, as shown below:

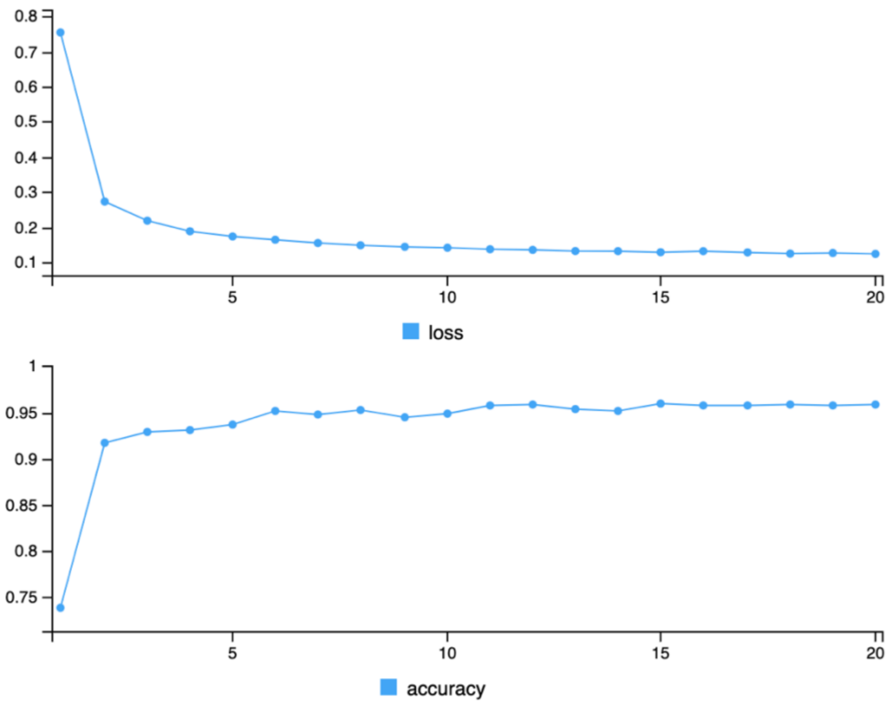


**Figure A.** Changes in loss function and accuracy over 20 epochs of training for the SNNN model to classify diabetes subgroups using NHANES-III data.

***Performance of the SNNN algorithm in NHANES-III and IV***

First, we compared the performance of using k-means clustering with variables available in MCPS to derive diabetes subgroups compared to the original cluster classification using the methods proposed by Ahlqvist et al. in the NHANES-IV validation sample, along with the performance of the classified subgroups using the SNNN model both in the training and validation samples. We observed decreased accuracy and concordance when using k-means clustering compared to the original variables, with marked worsening in the validation sample (**Table A**). In contrast, the SNNN model yielded improved accuracy and concordance, with similar performance in both the training and validation sample indicating the validity of the SNNN approach for reproducing diabetes subgroups proposed by Ahlqvist et al. using variables available in MCPS.

| Method | Accuracy (95%CI) | Kappa (95%CI) |
|---|---|---|
| Simple k-means clustering training sample | 0.785 (0.758-0.810) | 0.705 (0.669-0.741) |
| Simple k-means clustering validation sample | 0.737 (0.694-0.778) | 0.630 (0.577-0.683) |
| SNNN model validation sample | 0.924 (0.895-0.947) | 0.886 (0.829-0.944) |
| SNNN model training sample | 0.928 (0.901-0.943) | 0.899 (0.863-0.936) |

**Table A.** Performance and concordance of the k-means clustering compared to the SNNN approach for classifying diabetes subgroups as proposed by Ahlqvist et al. using training data in NHANES-III and validated in NHANES-IV for individuals with recently-diagnosed diabetes.

***Performance of the SNNN algorithm for specific diabetes subgroups***

Next, we assessed whether the SNNN model was adequate for identifying specific diabetes subgroups in the NHANES-IV validation sample. To this end, we compared sensitivity, specificity, positive and negative predictive values and balanced accuracy using the *caret* R package.We observed consistent performance across diabetes subgroups, with a slightly reduced accuracy for the MARD subgroup (**Table B**).

| Cluster | Sensitivity | Specificity | PPV | NPV | Balanced accuracy |
|---|---|---|---|---|---|
| SIDD | 99.72% | 90.32% | 97.5% | 98.8% | 95.02% |
| SIRD | 95.9% | 100.0% | 100.0% | 84.4% | 98.0% |
| MOD | 96.3% | 97.7% | 99.7% | 74.1% | 97.0% |
| MARD | 98.6% | 89.4% | 90.0% | 98.6% | 94.1% |

**Table B.** Diagnostic performance of the SNNN approach for classifying diabetes subgroups in NHANES-IV for individuals with recently-diagnosed diabetes. **Abbreviations:** MOD, Mild Obesity-related Diabetes; MARD, Mild Age-related Diabetes; SIDD, Severe Insulin-Deficient Diabetes; SIRD, Severe Insulin-Resistant Diabetes.

When assessing the clinical features of diabetes subgroups as reproduced by the SNNN algorithm we observed that the SIDD subgroup had the lowest HOMA2-B values, and the highest HbA1c levels compared to the rest of the subgroups. The SIRD subgroup had slightly increased HOMA2-B with elevated HOMA2-IR, BMI and WHtR values which are indicative of increased visceral adiposity. In contrast, the MARD subgroup was characterized by a milder phenotype with increased age at diabetes diagnosis, whilst the

MOD phenotype had the lowest age at diabetes diagnosis and the lowest HbA1c values, as seen in **Figure B**.

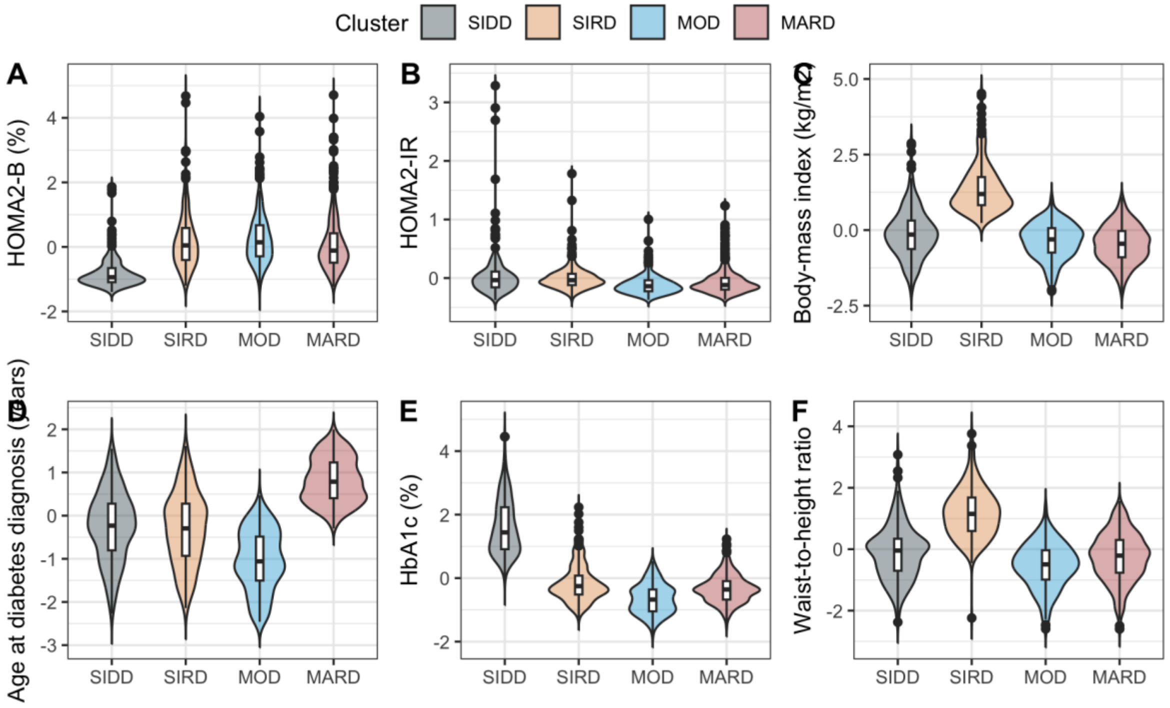


**Figure B.** Clinical characteristics of data-driven diabetes subgroups classified using SNNN algorithms in NHANES-III and NHANES-IV data. **Abbreviations:** MOD, Mild Obesity-related Diabetes; MARD, Mild Age-related Diabetes; SIDD, Severe Insulin-Deficient Diabetes; SIRD, Severe Insulin-Resistant Diabetes.

*Recalibration of SCORE2-Diabetes for fatal CVD outcomes*

To account for differences in baseline hazard and risk distribution between the for fatal outcomes compared to the original composite outcome of fatal and non-fatal outcomes, the SCORE2-Diabetes model was recalibrated for fatal cardiovascular disease (CVD) outcomes using a Cox proportional hazards framework. To this end, we let $h(t|\eta_i) = h_0(t)e^{\eta_i}$ denote the hazard function for subject $i$, where $h_0(t)$ is the baseline hazard and $\eta_i$ represents the original linear predictor for SCORE2-Diabetes estimated as the prognostic index. First, we estimated the recalibration slope for SCORE2-Diabetes by fitting a univariate Cox proportional hazard regression model using the linear predictor as the sole covariate, denoted by:

$$h(t|\eta_i) = h_0(t) * \exp(\beta_{recal}\eta_i),$$

where $\beta_{recal}$ denotes the calibration slope. This parameter quantifies the degree of over- or underestimation of risk by the original model and was estimated using partial maximum likelihood with the Efron approximation.

Next, we estimated the recalibrated baseline survival function with an individual with $\eta_i = 0$ from the fitted Cox model as:

$$\hat{S}_0(t) = \Pr(T > t \mid \eta_i = 0),$$

and evaluated the target prediction horizon $t_0$ =10 years, yielding $\hat{S}_0(t_0)$.

This transformation preserved the relative ranking of predicted risks while aligning the overall risk scale with the observed event rates in MCPS population. The recalibrated metrics for SCORE2-Diabetes are presented in **Table C** below:

| Metric | Estimate (95%CI |
|---|---|
| Recalibration slope ($\beta_{recal}$) | 0.4504 (0.4188-0.4821) |
| Baseline survival function $\hat{S}_0(t_0)$ | 0.979 (0.977-0.982) |

**Table C.** Recalibration metrics of SCORE2-Diabetes for fatal CVD in MCPS

Therefore, for each individual the recalibrated 10-year predicted probability of fatal CVD cab be computed as:

$$SCORE2 - Diabetes\ (fatal) = 1 - 0.979^{\exp(0.4504 * \eta_i)}.$$

Finally, we recalculated SCORE2-Diabetes for fatal CVD outcomes recalibrated for Mexican population using MCPS and compared its moderate calibration to the original model (**Figure C**). We observed better agreement between observed and predicted risks in the recalibrated model, indicating better risk estimates using this recalibrated version of SCORE2-Diabetes (fatal).

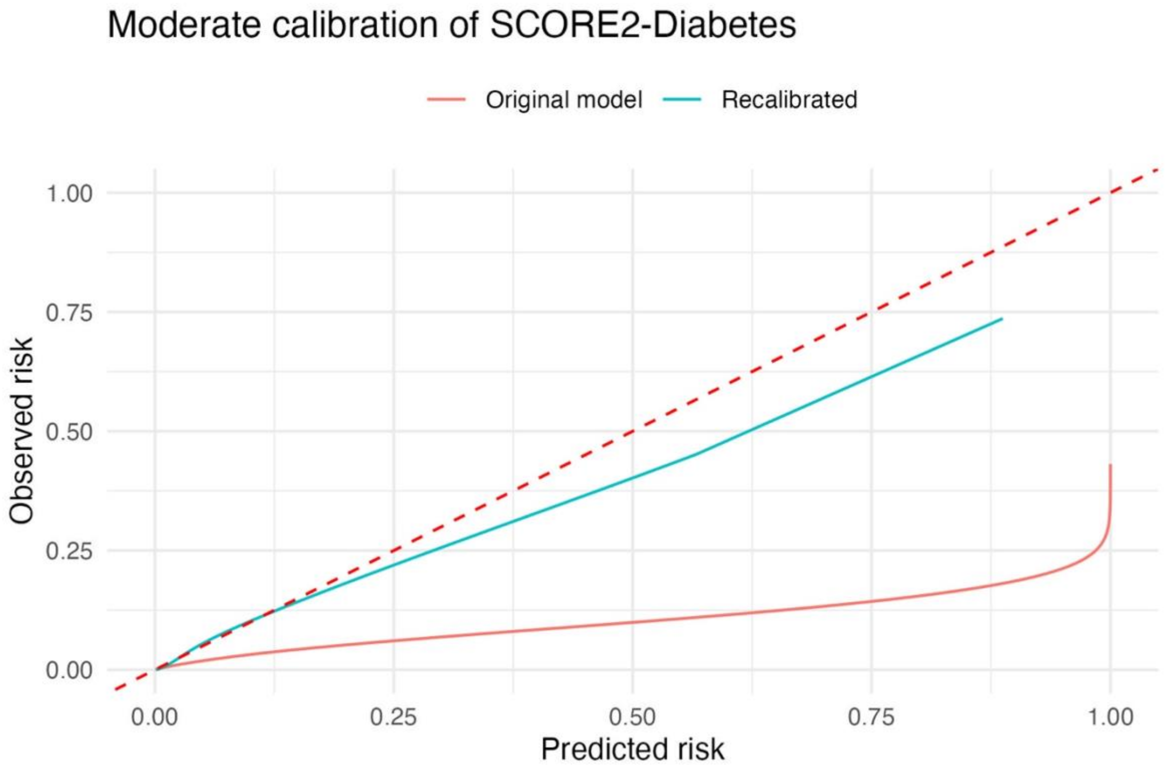


**Figure C.** Calibration curves of the original SCORE2-Diabetes model and its recalibration for risk of fatal CVD outcomes in MCPS.

**SUPPLEMENTARY TABLES**

**Supplementary Table 1**. Characteristics of participants of the Mexico City Prospective Study Cohort (n=159,517) and participants with diabetes at baseline (n=29,948).

| Variable | MCPS (n = 159,517) | | | MCPS with Diabetes (n = 29,948) | | |
|---|---|---|---|---|---|---|
| | All | Male | Female | All | Male | Female |
| | n = 159,517 | n = 52,579 (33%) | n = 106,938 (67%) | n = 29,948 | n = 9,988 (33.4%) | n = 19,960 (66.6%) |
| Age (years) | 53 (13) | 53 (13) | 52 (13) | 59 (12) | 59 (12) | 59 (12) |
| Follow-up (years) | 18.7 (5.0) | 18.2 (5.6) | 19.0 (4.6) | 16 (7) | 15 (7) | 17 (6) |
| Missing | 2,649 (1.7%) | 908 (1.7%) | 1,741 (1.6%) | 794 (2.7%) | 245 (2.5%) | 549 (2.8%) |
| Educational attainment (%) | | | | | | |
| University/College | 24,399 (15%) | 12,233 (23%) | 12,166 (11%) | 2,331 (7.8%) | 1,484 (15%) | 847 (4.2%) |
| High School | 38,529 (24%) | 13,336 (25%) | 25,193 (24%) | 4,676 (16%) | 2,057 (21%) | 2,619 (13%) |
| Elementary | 74,918 (47%) | 21,889 (42%) | 53,029 (50%) | 16,720 (56%) | 5,165 (52%) | 11,555 (58%) |
| Other | 21,587 (14%) | 5,100 (9.7%) | 16,487 (15%) | 6,198 (21%) | 1,279 (13%) | 4,919 (25%) |
| Missing | 84 (<0.1%) | 21 (<0.1%) | 63 (<0.1%) | 23 (<0.1%) | 3 (<0.1%) | 20 (0.1%) |
| Smoking (%) | 49,461 (31%) | 25,471 (48%) | 23,990 (22%) | 7,715 (26%) | 4,427 (44%) | 3,288 (16%) |
| Body mass index (kg/m²) | 29.0 (5.1) | 27.9 (4.3) | 29.5 (5.3) | 29.6 (5.5) | 28.2 (4.7) | 30.3 (5.7) |
| Missing | 2,129 (1.3%) | 869 (1.7%) | 1,260 (1.2%) | 666 (2.2%) | 281 (2.8%) | 385 (1.9%) |
| Systolic blood pressure (mmHg) | 128 (17) | 129 (16) | 127 (17) | 134 (18) | 133 (18) | 135 (18) |
| Missing | 320 (0.2%) | 168 (0.3%) | 152 (0.1%) | 51 (0.2%) | 29 (0.3%) | 22 (0.1%) |
| Glucose (mg/dl) | 99 (76) | 99 (78) | 99 (75) | 204 (119) | 204 (123) | 203 (117) |
| Missing | 6,684 (4.2%) | 2,409 (4.6%) | 4,275 (4.0%) | 1,032 (3.4%) | 380 (3.8%) | 652 (3.3%) |

| Variable | MCPS (n = 159,517) | | | MCPS with Diabetes (n = 29,948) | | |
|---|---|---|---|---|---|---|
| | **All** | **Male** | **Female** | **All** | **Male** | **Female** |
| | **n = 159,517** | **n = 52,579 (33%)** | **n = 106,938 (67%)** | **n = 29,948** | **n = 9,988 (33.4%)** | **n = 19,960 (66.6%)** |
| Glycated HbA1c (%) | 6.10 (1.71) | 6.10 (1.71) | 6.11 (1.71) | 8.83 (2.37) | 8.77 (2.39) | 8.86 (2.36) |
| Missing | 4,820 (3.0%) | 1,790 (3.4%) | 3,030 (2.8%) | 542 (1.8%) | 205 (2.1%) | 337 (1.7%) |
| Total cholesterol (mg/dl) | 164 (45) | 158 (44) | 166 (46) | 165 (49) | 156 (47) | 170 (50) |
| Missing | 6,684 (4.2%) | 2,409 (4.6%) | 4,275 (4.0%) | 1,032 (3.4%) | 380 (3.8%) | 652 (3.3%) |
| HDL cholesterol (mg/dl) | 37 (16) | 35 (15) | 39 (17) | 37 (18) | 35 (17) | 38 (18) |
| Missing | 6,684 (4.2%) | 2,409 (4.6%) | 4,275 (4.0%) | 1,032 (3.4%) | 380 (3.8%) | 652 (3.3%) |
| Non-HDL cholesterol (mg/dl) | 125 (41) | 122 (39) | 127 (41) | 127 (44) | 120 (43) | 130 (45) |
| Missing | 6,684 (4.2%) | 2,409 (4.6%) | 4,275 (4.0%) | 1,032 (3.4%) | 380 (3.8%) | 652 (3.3%) |
| Total triglycerides (mg/dl) | 136 (68) | 142 (70) | 133 (68) | 157 (82) | 156 (82) | 158 (82) |
| Missing | 6,684 (4.2%) | 2,409 (4.6%) | 4,275 (4.0%) | 1,032 (3.4%) | 380 (3.8%) | 652 (3.3%) |
| Apolipoprotein B (mg/dl) | 86 (41) | 85 (40) | 86 (42) | 88 (46) | 84 (45) | 89 (46) |
| Missing | 6,684 (4.2%) | 2,409 (4.6%) | 4,275 (4.0%) | 1,032 (3.4%) | 380 (3.8%) | 652 (3.3%) |
| Creatinine (mg/dl) | 0.73 (0.39) | 0.86 (0.46) | 0.66 (0.33) | 0.77 (0.61) | 0.91 (0.77) | 0.70 (0.50) |
| Missing | 6,684 (4.2%) | 2,409 (4.6%) | 4,275 (4.0%) | 1,032 (3.4%) | 380 (3.8%) | 652 (3.3%) |
| eGFR (ml/min/1.73m²) | 101 (17) | 100 (16) | 101 (17) | 95 (20) | 95 (21) | 95 (20) |
| Missing | 13,315 (8.3%) | 4,360 (8.3%) | 8,955 (8.4%) | 2,405 (8.0%) | 806 (8.1%) | 1,599 (8.0%) |

Mean (SD) for continuous variables, n (%) for categorical variables.

**Supplementary Table 2**. Characteristics of participants of the Mexico City Prospective Study Cohort with diabetes and without cardiovascular disease or missing data for cluster classification at baseline (n=24,943).

| Variable | All<br>n = 24,943 | Male<br>n = 8,188 (32.8%) | Female<br>n = 16,755 (67.2%) |
|---|---|---|---|
| Age (years) | 58 (12) | 58 (12) | 58 (12) |
| Follow-up (years) | 16 (6) | 16 (7) | 17 (6) |
| Educational attainment (%) | | | |
| University/College | 1,951 (7.8%) | 1,242 (15%) | 709 (4.2%) |
| High School | 4,003 (16%) | 1,737 (21%) | 2,266 (14%) |
| Elementary | 13,969 (56%) | 4,218 (52%) | 9,751 (58%) |
| Other | 5,000 (20%) | 990 (12%) | 4,010 (24%) |
| Missing | 20 (<0.1%) | 1 (<0.1%) | 19 (0.1%) |
| Smoking (%) | 6,539 (26%) | 3,709 (45%) | 2,830 (17%) |
| Body mass index (kg/m²) | 29.7 (5.4) | 28.2 (4.6) | 30.4 (5.7) |
| Systolic blood pressure (mmHg) | 134 (18) | 133 (17) | 134 (18) |
| Glucose (mg/dl) | 207 (117) | 208 (122) | 206 (114) |
| Glycated HbA1c (%) | 8.86 (2.37) | 8.81 (2.39) | 8.88 (2.36) |
| Total cholesterol (mg/dl) | 169 (40) | 160 (38) | 174 (40) |
| HDL cholesterol (mg/dl) | 39 (8) | 36 (7) | 40 (8) |
| Non-HDL cholesterol (mg/dl) | 130 (37) | 123 (35) | 133 (37) |
| Total triglycerides (mg/dl) | 164 (70) | 163 (71) | 164 (70) |
| Apolipoprotein B (mg/dl) | 92 (23) | 88 (22) | 94 (23) |
| Creatinine (mg/dl) | 0.80 (0.56) | 0.94 (0.74) | 0.73 (0.43) |
| eGFR (ml/min/1.73m²) | 96 (20) | 96 (20) | 95 (20) |

Mean (SD) for continuous variables, n (%) for categorical variables.

**Supplementary Table 3**. Fatal cardiovascular events in participants of the Mexico City Prospective Study Cohort overall and in those with diabetes and without cardiovascular disease or missing data for cluster classification at baseline.

| **Variable** | **MCPS (n = 159,517)** | | | **MCPS with Diabetes (n = 24,943)** | | |
|---|---|---|---|---|---|---|
| | **All** | **Male** | **Female** | **All** | **Male** | **Female** |
| | **n = 159,517 (100%)** | **n = 52,579 (33%)** | **n = 106,938 (67%)** | **n = 24,943 (100%)** | **n = 8,188 (32.8%)** | **n = 16,755 (67.2%)** |
| **Narrow definition**[1] | | | | | | |
| Total cardiovascular events | 7,565 (4.8%) | 3,278 (6.3%) | 4,287 (4.1%) | 2,218 (8.9%) | 898 (11%) | 1,320 (7.9%) |
| Cerebrovascular events | 1,311 (0.8%) | 472 (0.9%) | 839 (0.8%) | 368 (1.5%) | 131 (1.6%) | 237 (1.4%) |
| Ischemic cardiac events | 6,254 (4.0%) | 2,806 (5.4%) | 3,448 (3.3%) | 1,850 (7.4%) | 767 (9.4%) | 1,083 (6.5%) |
| **Broad definition**[2] | | | | | | |
| Total cardiovascular events | 8,946 (5.7%) | 3,763 (7.3%) | 5,183 (4.9%) | 2,583 (10%) | 1,010 (12%) | 1,573 (9.4%) |
| Cerebrovascular events | 1,311 (0.8%) | 472 (0.9%) | 839 (0.8%) | 368 (1.5%) | 131 (1.6%) | 237 (1.4%) |
| Ischemic cardiac events | 6,254 (4.0%) | 2,806 (5.4%) | 3,448 (3.3%) | 1,850 (7.4%) | 767 (9.4%) | 1,083 (6.5%) |
| *Other cardiac events* | | | | | | |
| Arrythmias | 113 (<0.1%) | 29 (<0.1%) | 84 (<0.1%) | 28 (0.1%) | 6 (<0.1%) | 22 (0.1%) |
| Heart failure | 455 (0.3%) | 156 (0.3%) | 299 (0.3%) | 121 (0.5%) | 40 (0.5%) | 81 (0.5%) |
| Hypertensive disease | 768 (0.5%) | 272 (0.5%) | 496 (0.5%) | 196 (0.8%) | 54 (0.7%) | 142 (0.8%) |
| Other | 23 (<0.1%) | 16 (<0.1%) | 7 (<0.1%) | 7 (<0.1%) | 6 (<0.1%) | 1 (<0.1%) |
| *Other vascular events* | | | | | | |
| Peripheral arterial disease | 66 (<0.1%) | 29 (<0.1%) | 37 (<0.1%) | 25 (0.1%) | 8 (<0.1%) | 17 (0.1%) |

[1]ICD-10: I20-I25,I60-I69.

[2]ICD-10: I20-I25,I10-I15,I46-I52,I60-I69,I170-I179, R96,E105,E115,E145.

**Supplementary Table 4**. Fatal cardiovascular events in participants of the Mexico City Prospective Study Cohort in those with diabetes and without cardiovascular disease or missing data for cluster classification at baseline, classified per cluster. **Abbreviations:** MOD, Mild Obesity-related Diabetes; MARD, Mild Age-related Diabetes; SIDD, Severe Insulin-Deficient Diabetes; SIRD, Severe Insulin-Resistant Diabetes.

| Variable | **All** n = 24,943 | **SIDD** n = 12,466 (50%) | **MARD** n = 4,409 (17.7%) | **SIRD** n = 3,512 (14.1%) | **MOD** n = 4,556 (18.3%) |
|---|---|---|---|---|---|
| | | | **Narrow definition[1]** | | |
| Total cardiovascular events | 2,218 (8.9%) | 1,178 (9.4%) | 563 (13%) | 201 (5.7%) | 276 (6.1%) |
| Cerebrovascular events | 368 (1.5%) | 188 (1.5%) | 104 (2.4%) | 38 (1.1%) | 38 (0.8%) |
| Ischemic cardiac events | 1,850 (7.4%) | 990 (7.9%) | 459 (10%) | 163 (4.6%) | 238 (5.2%) |
| | | | **Broad definition[2]** | | |
| Total cardiovascular events | 2,583 (10%) | 1,354 (11%) | 663 (15%) | 256 (7.3%) | 310 (6.8%) |
| Cerebrovascular events | 368 (1.5%) | 188 (1.5%) | 104 (2.4%) | 38 (1.1%) | 38 (0.8%) |
| Ischemic cardiac events | 1,850 (7.4%) | 990 (7.9%) | 459 (10%) | 163 (4.6%) | 238 (5.2%) |
| *Other cardiac events* | | | | | |
| Arrythmias | 28 (0.1%) | 12 (<0.1%) | 10 (0.2%) | 3 (<0.1%) | 3 (<0.1%) |
| Heart failure | 121 (0.5%) | 54 (0.4%) | 36 (0.8%) | 23 (0.7%) | 8 (0.2%) |
| Hypertensive disease | 196 (0.8%) | 99 (0.8%) | 53 (1.2%) | 29 (0.8%) | 15 (0.3%) |
| Other | 7 (<0.1%) | 4 (<0.1%) | 0 (0%) | 0 (0%) | 3 (<0.1%) |
| *Other vascular events* | | | | | |
| Peripheral arterial disease | 25 (0.1%) | 13 (0.1%) | 10 (0.2%) | 2 (<0.1%) | 0 (0%) |

[1]ICD-10: I20-I25,I60-I69.

[2]ICD-10: I20-I25,I10-I15,I46-I52,I60-I69,I170-I179, R96,E105,E115,E145.

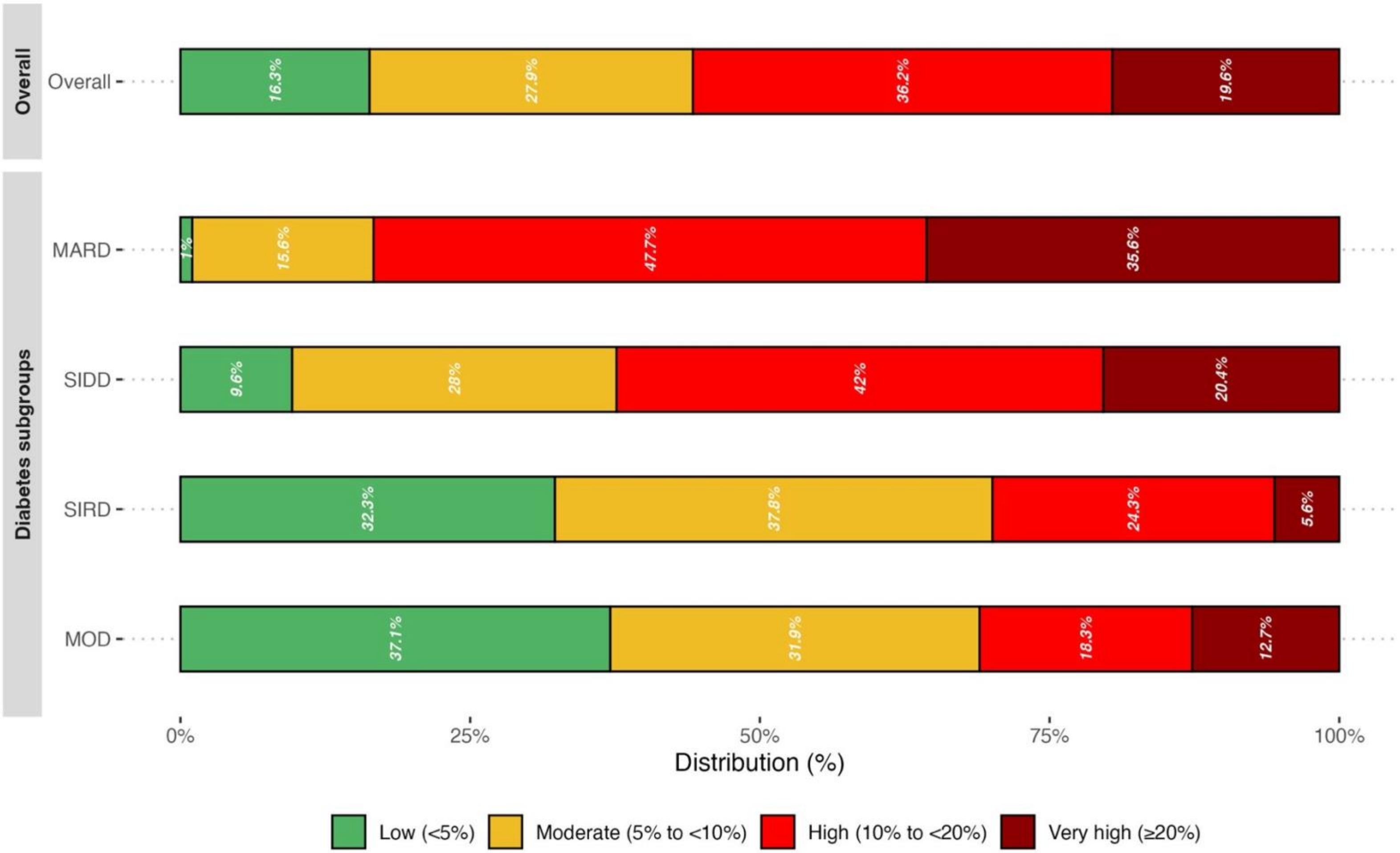


**Supplementary Figure 1.** Distribution of the SCORE2-Diabetes risk categories overall and across diabetes subgroups in MCPS using the original version of SCORE2-Diabetes.

**Abbreviations:** MOD, Mild Obesity-related Diabetes; MARD, Mild Age-related Diabetes; SIDD, Severe Insulin-Deficient Diabetes; SIRD, Severe Insulin-Resistant Diabetes.

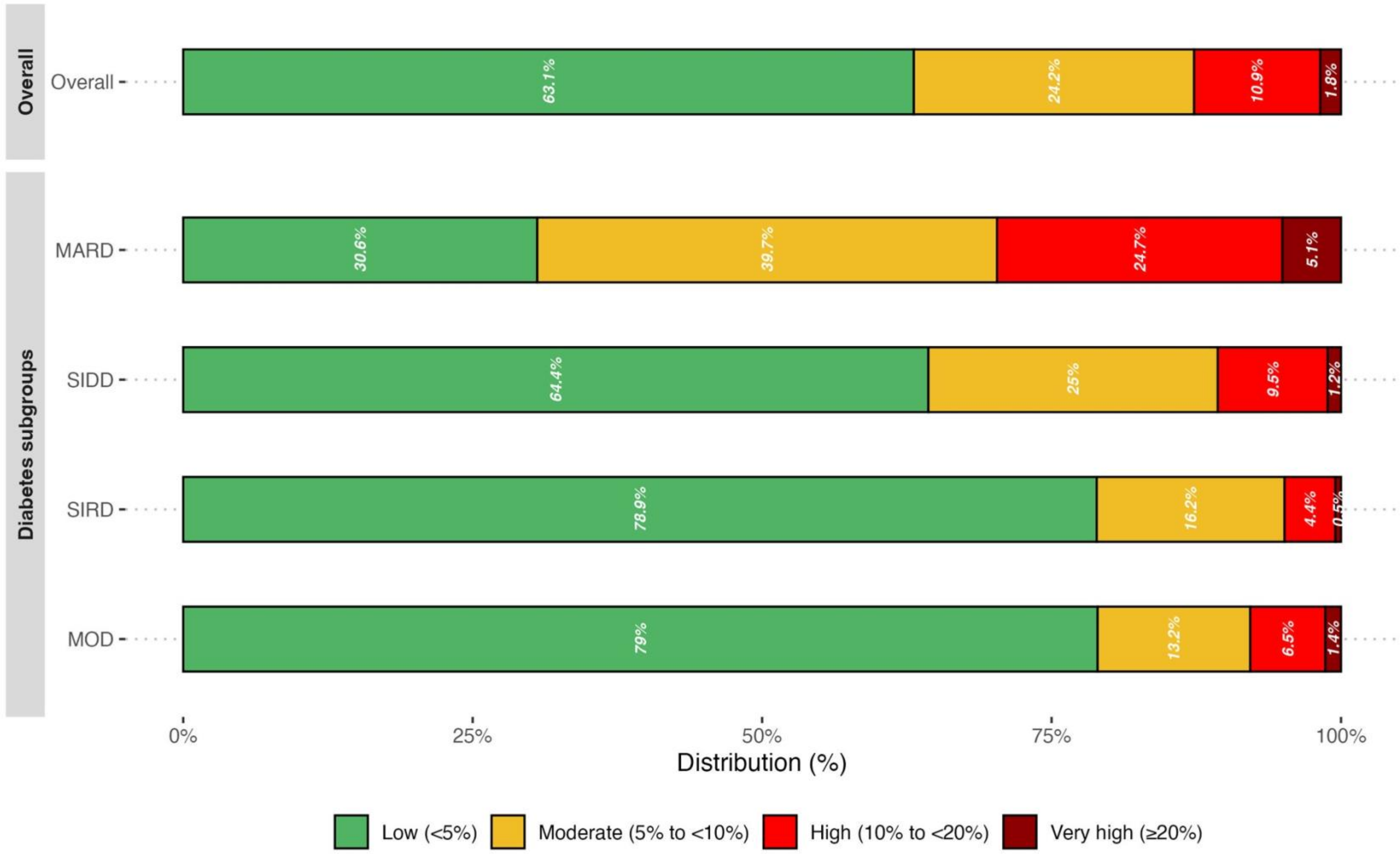


**Supplementary Figure 2.** Distribution of the SCORE2-Diabetes risk categories overall and across diabetes subgroups in MCPS using the recalibrated version of SCORE2-Diabetes for fatal CVD outcomes.

**Abbreviations:** MOD, Mild Obesity-related Diabetes; MARD, Mild Age-related Diabetes; SIDD, Severe Insulin-Deficient Diabetes; SIRD, Severe Insulin-Resistant Diabetes.
